## Additional File 1 for "The experience of European hospital-based health care workers on following infection prevention and control procedures for COVID-19"

**ADDITIONAL FILE 1 – SURVEY TOOLS**

**1. SURVEY TOOL ROUND 1**

| I understand that no personal data will be collected and that my participation in this study is completely voluntary. | Yes/ no* |
| --- | --- |
| I consent to take part in this study. | Yes/ no* |
| Do you currently provide direct medical care to patients, or do you expect to provide direct medical care to patients during the upcoming four weeks? | Yes/ no* / not sure  * not eligible for participation. |
| Age | Number in years |
| Gender | Female, male, prefer not to say |
| Hospital type | Academic hospital  Teaching hospital  General hospital  Unsure (+ open text field) |
| Current marital status | A. Living alone:  Single (never married)  Divorced  Separated  Widowed  Other (namely)  B. Living with others:  Married / Civil partnership  Other (namely)  C. Prefer not to say |
| Number of children currently in household, under the age of 17 years | 1, 2, 3, 4, 5+  None  Prefer not to say |
| Do you have caring responsibilities for any adults, including those with disabilities or those over the age of 70 years? | Yes  No  Prefer not to say |
| Current job role | Senior medical doctor  Junior medical doctor  Nurse  Allied health professional  Other (+ namely) |
| In your current job role as healthcare worker, how frequently do you have direct patient contact? | 1) daily 2) more than one day per week, 3) less than five days per month, 4) rarely, 5) don’t know |
| Type of job role | Fulltime/ part time/ casual or locum staff / retired / prefer not to say |
| Length of time since completed medical training | Number in years |

I. Have you previously worked in a clinical setting during an acute respiratory epidemic or pandemic, for example, SARS (2002), MERS Co-V (2012), H1N1 (2009)?

II. In a clinical setting, did you personally care for patients with suspected or confirmed infection caused by a novel respiratory pathogen for example, SARS, MERS Co-V, H1N1?

III. Has a patient with suspected or confirmed COVID-19 attended the hospital in which you work?

IV. Have you personally cared for a patient with suspected or confirmed COVID-19 infection?

**IF YES TO IV:**

IVa. What was your most recent type of contact with a suspected/confirmed COVID-19 case?

a. Close contact: directly caring for a suspected/confirmed patient or being within a 1-2m radius of a suspected/confirmed patient

b. Healthcare facility contact: no direct contact with suspected/confirmed COVID-19 case, however worked in the same facility

c. Unknown / unsure

IF a.: Did this contact include an aerosol generating procedure? For example tracheal intubation, non-invasive ventilation, bronchoscopy, cardiopulmonary resuscitation.

IVb. Did this most recent patient have confirmed or suspected COVID-19?

IVc. What kind of infection prevention procedures did you use during your most recent contact with a suspected/confirmed COVID-19 case?

- Hand hygiene	Yes/no/not sure
- N95 respirator (FFP2 or equivalent)	Yes/no/not sure
  - If Yes:	PP1, PP2, PP3, other
- Other types of medical mask	Yes/no/not sure
  - If Yes:	Which one?
- Fluid-resistant gown	Yes/no/not sure
- Disposable plastic apron	Yes/no/not sure
- Gloves	Yes/no/not sure
- Full body suit	Yes/no/not sure
- Eye protection (i.e. goggles or face shield)	Yes/no/not sure
- Single use equipment	Yes/no/not sure
- Other	Open text field

**IF NO TO IV:**

IVd. When providing direct medical care to suspected or confirmed COVID-19 cases, excluding during aerosol-generating procedures, which of the following procedures are **currently recommended** in your hospital for preventing transmission?

- Hand hygiene	Yes/no/not sure
- N95 respirator (FFP2 or equivalent)	Yes/no/not sure
- Other types of medical mask	Yes/no/not sure
- Fluid-resistant gown	Yes/no/not sure
- Disposable plastic apron	Yes/no/not sure
- Gloves	Yes/no/not sure
- Full body suit	Yes/no/not sure
- Eye protection (i.e. goggles or face shield)	Yes/no/not sure
- Single use equipment	Yes/no/not sure
- Other	Open text field

V. In my hospital, infection prevention and control procedures are different for SUSPECTED and CONFIRMED COVID-19 cases.

The following questions relate to your experience of managing patients in the healthcare setting where you work. Please think about your experience **over the past week** when responding to these questions.

*Response options: 7 point Likert scale: strongly disagree, disagree, somewhat disagree, neutral, somewhat agree, agree, strongly agree.*

1. I am confident that the hospital where I work can manage current patient demand related to COVID-19.

2. I am confident that the hospital where I work can continue to manage patient demand related to COVID-19 over the next 3 months.

3. I have received general training for infection, prevention and control procedures for communicable diseases.

4. I have received sufficient training in the infection prevention and control practices specifically for COVID-19.

5. I am confident in my ability to correctly don and doff personal protective equipment to prevent transmission of COVID-19 to others and myself.

6. I am confident that I am able to follow recommended procedures related to personal protective equipment (PPE) for COVID-19 e.g. appropriate use and disposal of gloves, apron and surgical mask.

7. I consider that the implementation of protective procedures at work are effective to prevent the spread of COVID-19 in my hospital.

8. Following the infection prevention and control recommendations will protect me from becoming ill with COVID-19.

9. Following recommended infection prevention and control procedures adds significant additional strain to my workload.

10. I will always use the recommended personal protective equipment (medical mask, eye protection, gown and gloves) when taking care of patients with suspected or confirmed COVID-19 when I have access to these.

11. There are clear policies and protocols in my hospital for everyone to follow related to infection prevention and control procedures for COVID-19.

12. I can easily access personal protective equipment (PPE) in line with standard infection control precautions, such as gloves, apron and masks, for COVID-19 in the hospital where I work.

13. In my hospital there are dedicated isolation facilities for patients with COVID-19.

14. I know who to contact if I have trouble with PPE, or if I have a body fluid exposure/ unprotected contact with a confirmed case of COVID-19.

15. The hospital where I work receives good support from national/ regional/ local public health authorities, who provide guidance and training on how to manage COVID-19.

16. Most of my colleagues regularly follow infection, prevention and control measures (for example, regular hand washing, use of personal protective equipment, proper disposal of equipment).

17. It is expected that in my role as a healthcare professional that I will follow infection prevention and control measures.

18. I am encouraged and supported by senior medical/nurse staff to apply recommended infection prevention and control measures.

19. I am concerned about the risk to myself of becoming ill with COVID-19.

20. I am concerned about the risk to my family related to COVID-19 as a result of my job role.

21. I am afraid of looking after patients who are ill with COVID-19.

22. I accept that the risk of getting COVID-19 is part of my job.

23. Whether I get infected with COVID-19 is within my control.

24. The health facility where I work is competent to manage COVID-19.*

25. The health facility where I work are being honest with staff when managing COVID-19.*

26. The health facility where I work would act in the best interests of its staff when managing COVID-19.*

**Note: these 3 questions are combined in the analysis to create a single “trust” score.*

27. During your last clinical shift, what was the availability of the following materials:

*(response options: none available, limited supply, moderate supply, full supply)*

- Hand alcohol
- N95 respirator (FFP2 or equivalent)
- Surgical mask
- Fluid-resistant gown
- Disposable apron
- Disposable gloves
- Full body suit
- Eye protection (i.e. goggles or face shield)

WHO-5 Well-Being Index

Over the last two weeks:

*(response options: all of the time; most of the time; more than half of the time; less than half of the time; some of the time; at no time)*

28. I have felt cheerful and in good spirits.

29. I have felt calm and relaxed.

30. I have felt active and vigorous.

31. I woke up feeling fresh and rested.

32. My daily life has been filled with things that interest me.

**2. SURVEY TOOL ROUND 2**

| I understand that no personal data will be collected and that my participation in this study is completely voluntary. | Yes/ no* |
| --- | --- |
| I consent to take part in this study. | Yes/ no |
| Screening question: do you currently provide direct medical care to hospital patients, or do you expect to provide direct medical care to hospital patients during the upcoming 4 weeks? | Yes/ no*  * Not eligible for participation |
| What is your age? | Number in years |
| What is your gender? | Female, male, prefer not to say |
| In which country do you currently work? | Standardized dropdown field |
| What hospital type are you currently working in? | Academic hospital  Non-academic hospital  Unsure |
| What is your current* living situation?  **Please select the option that was best applicable during the majority of the time during the past 2 weeks.* | Living alone  Living with others  Prefer not to say |
| Do you have caring responsibilities* for any adults, including those with disabilities or those over the age of 70 years?  **With caring responsibilities we mean if you regularly look after someone, which is not part of your medical profession.* | Yes  No  Prefer not to say |
| What is your current job role*?  **With current job role we mean the role in which you currently provide medical care to patients.* | Student doctor (undergraduate)  Junior medical doctor  Senior medical doctor  Retired doctor (returned to practice)  Student nurse  Junior nurse  Senior nurse  Retired nurse (returned to practice)  Student allied health professional (i.e. physician assistant)  Junior allied health professional  Senior allied health professional  Retired allied health professional  Other |
| In what medical specialty do you currently provide medical care to patients?*  **Select the option that best applies during your current work in the COVID-19 pandemic.* | Acute care medicine (i.e ICU, anaesthesiology)  Internal medicine  Surgery  Paediatrics  Other (i.e. gynaecology, neurology) |
| In your current job role, how frequently do you have direct patient contact*?  **Select the option that best applies during your current medical work in the COVID-19 pandemic* | Daily  More than one day per week  Less than five days per month  Rarely  Don’t know |

I. Have you previously worked in a clinical setting during an acute respiratory epidemic or pandemic, for example, SARS (2002), MERS Co-V (2012), H1N1 (2009)? Yes/no/unsure

IF “YES” OR “UNSURE” TO I:

II. In a clinical setting, did you personally care for patients with suspected or confirmed infection caused by a previous novel respiratory pathogen for example, SARS (2002), MERS Co-V (2012), H1N1 (2009)? Yes/no/unsure

III. Have you personally provided direct (medical) care to a hospital patient with suspected or confirmed COVID-19 infection? Yes/no

IF YES TO III:

IV. Did your most recent medical contact with a suspected or confirmed COVID-19 patient include an aerosol generating procedure? For example tracheal intubation, non-invasive ventilation, bronchoscopy, tracheotomy, manual ventilation before intubation, cardiopulmonary resuscitation. Yes/no/unsure

V. At your most recent contact with a suspected/confirmed COVID-19 patient, what kind of infection prevention procedures did you use?

- Hand hygiene	Yes/no/unsure
- N95 respirator (FFP2 or equivalent)	Yes/no/unsure
- Other type of face mask (i.e. surgical mask)	Yes/no/unsure
- Eye protection (i.e. goggles, face shield)	Yes/no/unsure
- Fluid-resistant long-sleeved gown	Yes/no/unsure
- Disposable apron	Yes/no/unsure
- Full-body suit	Yes/no/unsure
- Gloves	Yes/no/unsure
- If applicable: other (namely)	Yes/no/unsure

The following questions relate to your experience of managing patients in the healthcare setting where you work. Please think about your experience **over the past week** when responding to these questions.

*Response options: 7 point Likert scale: strongly disagree, disagree, somewhat disagree, neutral, somewhat agree, agree, strongly agree.*

1. I am confident that the hospital where I work can manage current patient demand related to COVID-19

2. I have received general training for infection, prevention and control procedures for communicable diseases

3. I have received sufficient training in the infection prevention and control practices specifically for COVID-19

4. I am confident in my ability to correctly don and doff personal protective equipment (PPE) to prevent transmission of COVID-19 to others and myself.

5. I am confident that I am able to follow recommended procedures related to personal protective equipment (PPE) for COVID-19 e.g. appropriate use and disposal of gloves, apron and fluid resistant surgical mask.

6. I believe that the protective procedures at work are sufficiently effective to prevent the spread of COVID-19 in my hospital.

7. Following the infection prevention and control recommendations will protect me from becoming ill with COVID-19.

8. Following recommended infection prevention and control procedures adds significant additional strain to my workload.

9. I intend to always use the recommended personal protective equipment (medical mask, eye protection, gown and gloves) when taking care of patients with suspected or confirmed COVID-19 when I have access to these.

******If you are currently not providing direct care to COVID-19 patients, please think of a future possible situation where you would, when answering this question*

10. There are clear policies and protocols in my hospital for everyone to follow related to infection prevention and control procedures for COVID-19

11. I can easily access personal protective equipment (PPE) in line with standard infection control precautions, such as gloves, apron and masks, for COVID-19 in the hospital where I work

12. During your last clinical shift, what was the availability of the following materials?

*(response options: none available, limited supply, moderate supply, full supply)*

- Hand alcohol
- N95 respirators (FFP2 or equivalent)
- Surgical masks
- Fluid-resistant long-sleeved gown
- Disposable aprons
- Disposable gloves
- Eye protection (i.e. goggles or face shields)

13. In my hospital there are dedicated isolation facilities for patients with COVID-19.

14. There is a designated person in my health facility to contact if I have trouble with PPE, or if I have a body fluid exposure/ unprotected contact with a confirmed case of COVID-19.

15. The hospital where I work receives good support from national/ regional/ local public health authorities, who provide guidance and training on how to manage COVID-19.

16. Most of my colleagues regularly follow infection, prevention and control measures (for example, regular hand washing, use of personal protective equipment, proper disposal of equipment).

17. It is expected that in my role as a healthcare professional I will follow infection prevention and control measures.

18. I am encouraged and supported by senior medical/nurse staff to apply recommended infection prevention and control measures.

19. I am concerned about the risk to myself of becoming ill with COVID-19.

20. I am concerned about the risk to my family related to COVID-19 as a result of my job role.

21. I am afraid of looking after patients who are ill with COVID-19.

22. I accept that the risk of getting COVID-19 is part of my job.

23. Whether I get infected with COVID-19 is within my control.

WHO-5 Well-Being Index

Over the last two weeks:

*(response options: all of the time; most of the time; more than half of the time; less than half of the time; some of the time; at no time)*

24. I have felt cheerful and in good spirits

25. I have felt calm and relaxed

26. I have felt active and vigorous

27. I woke up feeling fresh and rested

28. My daily life has been filled with things that interest me

29. The health facility where I work is competent to manage COVID-19*

30. The health facility where I work are being honest with staff when managing COVID-19*

31. The health facility where I work would act in the best interests of its staff when managing COVID-19.*

**Note: these 3 questions are combined in the analysis to create a single “trust” score.*
