## Additional File 2 for "The experience of European hospital-based health care workers on following infection prevention and control procedures for COVID-19"

| **eTable 1.** Country of work of responding hospital healthcare workers (HCWs). | | |
| --- | --- | --- |
|  | **Round 1**  **N = 190 (%)** | **Round 2**  **N = 2099 (%)** |
| Albania | - | 7 (0.3) |
| Austria | 1 (0.5) | 3 (0.1) |
| Belarus | - | 2 (0.1) |
| Belgium | 4 (2.1) | 43 (2.0) |
| Bosnia and Herzegovina | - | 2 (0.1) |
| Bulgaria | 2 (1.2) | 3 (0.1) |
| Croatia | 8 (4.2) | 2 (0.1) |
| Cyprus | 1 (0.5) | 9 (0.4) |
| Czech Republic | 1 (0.5) | 3 (0.1) |
| Denmark | 3 (1.6) | 4 (0.2) |
| Estonia | 3 (1.6) | 29 (1,4) |
| Finland | - | 1 (0.1) |
| France | 8 (4.2) | 166 (7.9) |
| Germany | 6 (3.2) | 159 (7.6) |
| Greece | 23 (12) | 42 (2.0) |
| Hungary | 6 (3.2) | 7 (0.3) |
| Ireland | 2 (1.1) | 5 (0.2) |
| Israel | - | 4 (0.2) |
| Italy | 7 (3.7) | 54 (2.6) |
| Latvia | 3 (1.6) | 24 (1.1) |
| Lithuania | 1 (0.5) | - |
| Luxembourg | - | 1 (0.1) |
| Macedonia | 3 (1.6) | 3 (0.1) |
| Malta | - | 147 (7.0) |
| Montenegro | 1 (0.5) | - |
| Netherlands | 13 (6.8) | 242 (11.5) |
| Norway | - | 183 (8.7) |
| Poland |  | 13 (0.6) |
| Portugal | 18 (9.5) | 712 (33.9) |
| Republic of Kosovo | - | 2 (0.1) |
| Romania | 9 (4.7) | 8 (0.4) |
| Serbia | 1 (0.5) | 33 (1.6) |
| Slovakia | - | 1 (0.1) |
| Slovenia | 1 (0.5) | 1 (0.1) |
| Spain | 55 (28.9) | 98 (4.7) |
| Sweden | - | 2 (0.1) |
| Switzerland | - | 12 (0.6) |
| Turkey | 1 (0.5) | 9 (0.4) |
| Ukraine | - | 2 (0.1) |
| United Kingdom | 8 (4.2) | 61 (2.9) |
| Unknown | 1 (0.5) | - |

| **eTable 2.** Round 1 survey: comparison overall invited group of health care workers and those who responded. | | | |
| --- | --- | --- | --- |
|  | *Recipients survey*  *N = 2436 (%)* | *Respondents*  *N = 190 (%)* | *P value*^1^ |
| **Female** | 921 (37.8) | 84 (42.6) | NS |
| **Region^2^**  Eastern Europe  Northern Europe  Southern Europe  Western Europe  Unknown | 384 (15.8)  248 (10.2)  1083 (44.5)  715 (29.4)  6 (0.2) | 18 (9.5)  20 (10.5)  119 (62.6)  32 (16.8)  1 (0.5) | <.05 |
| **Medical specialty**  Acute care (anaesthesiology, ER, ICU)  Internal medicine  *Of which infectious diseases*  Public health & research  Surgery  Other  Unknown | 1068 (43.8)  885 (36.3)  *586 (24.1)*  196 (8.0)  181 (7.4)  69 (2.8)  37 (1.5) | 69 (36.3)  76 (40.0)  *36 (18.9)*  13 (6.8)  14 (7.4)  11 (5.8)  7 (3.7) | <.05 |
| ER, emergency room; ICU, intensive care unit; NS, non-significant ^1^ *C*omparison characteristics of responding sample to expected proportions based on overall survey recipient group, *P* value from Chi-square goodness of fit test.  ^2^ Sub division of Europe adapted from the United Nations; for the current study, Cyprus, Israel and Turkey were categorized as Southern Europe [35]. | | | |

**eFigure 1.** Self-reported availability of personal protective equipment during most recent clinical shift.

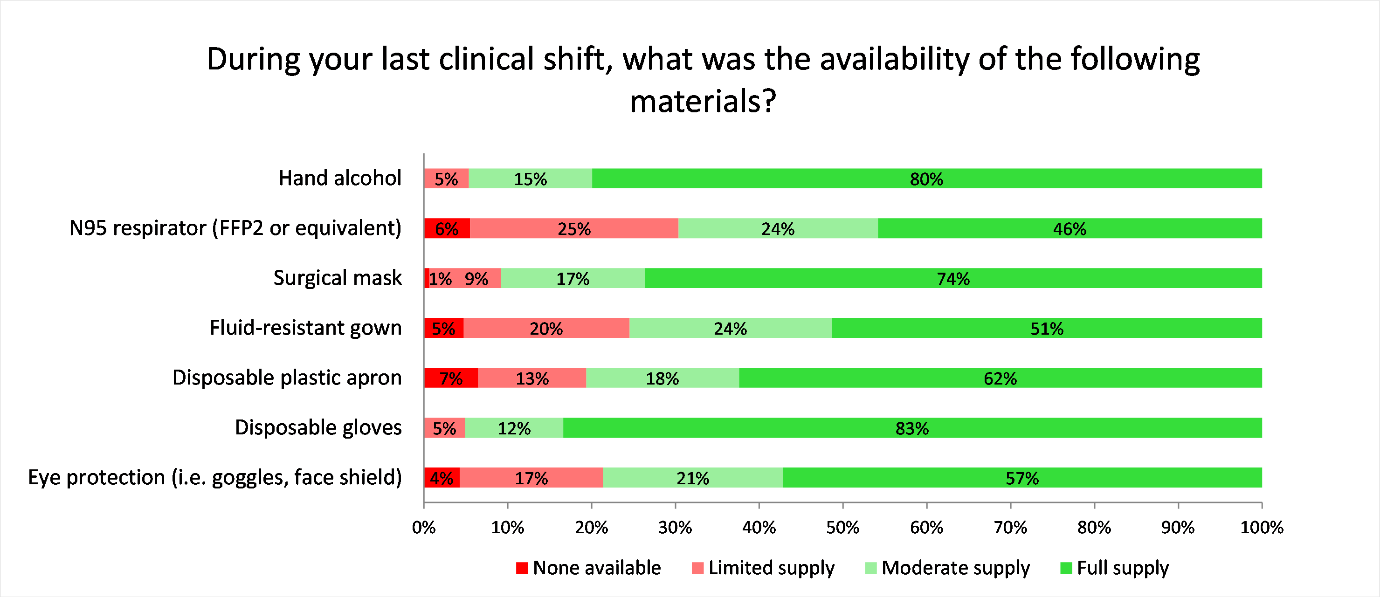

| **eTable 3.** Multivariable logistic regression for the association between gender and a WHO-5 Well-being Index below 50 points, in hospital healthcare workers during the COVID-19 pandemic. | | | | |
| --- | --- | --- | --- | --- |
|  | **aOR** | **95% CI** | | ***P value*** |
| Age (in years) | 1.0 | 1.0 | 1.0 | .03 |
| Female gender | 1.5 | 1.2 | 1.8 | <.001 |
| Living alone | 1.2 | 0.9 | 1.6 | NS |
| Job role  Other  Junior nurse  Senior nurse  Junior medical doctor  Senior medical doctor  Junior allied health professional  Senior allied health professional | ref  1.0  1.0  1.0  1.1  1.0  1.5 | ref  0.7  0.7  0.6  0.7  0.5  0.9 | ref  1.6  1.5  1.5  1.5  2.3  2.8 | -  NS  NS  NS  NS  NS  NS |
| Academic hospital | 0.9 | 0.8 | 1.1 | NS |
| Region^1^  Western Europe  Eastern Europe  Southern Europe  Northern Europe | ref  3.6  2.3  1.8 | ref  2.0  1.8  1.3 | ref  6.6  2.9  2.4 | -  <.001  <.001  <.001 |
| Providing direct COVID-19 patient care | 1.3 | 1.0 | 1.6 | .04 |
| CI, confidence interval; COVID-19, coronavirus disease 2019; HCW, healthcare worker; OR, odds ratio; WHO, World Health Organization  ^1^ Sub division of Europe adapted from the United Nations; for the current study, Cyprus, Israel and Turkey were categorized as Southern Europe [35]. | | | | |
